# Modulation of Neural Networks and Symptom Correlated in Fibromyalgia: A Randomized Double-blind Factorial Explanatory Clinical Trial of Home-Based Transcranial Direct Current Stimulation

**DOI:** 10.1101/2023.07.05.23292267

**Authors:** Rael Lopes Alves, Maxciel Zortea, Paul Vicuña Serrano, Vani dos Santos Laranjeira, Betina Franceschini Tocchetto, Leticia Ramalho, Camila Fernanda da Silveira Alves, Rafaela Brugnera Tomedi, Rodrigo Pereira de Almeida, Samara Machado Bruck, Liciane Medeiros, Paulo R. S. Sanches, Danton P. Silva, Iraci Lucena da Silva Torres, Felipe Fregni, Wolnei Caumo

## Abstract

Transcranial direct current stimulation (tDCS) might modulate neural activity and promote neural plasticity. This factorial randomized clinical trial compared a-tDCS on the left dorsolateral prefrontal cortex (l-DLPFC) or sham (s-tDCS), and a-tDCS or s-tDCS on the primary motor cortex (M1) in the connectivity analyses in eight regions of interest (ROIs) across eight resting-state electroencephalography (EEG) frequencies. We included 48 women with fibromyalgia, aged 30 to 65, randomly assigned to 2:1:2:1 to receive 20 sessions during 20 minutes of a-tDCS 2mA or s-tDCS at home, over l-DLPFC or M1, respectively. EEG recordings were obtained before and after treatment with eyes open (EO) and eyes closed (EC). In the EC condition, comparing pre to post-treatment, the a-tDCS on l-DLPFC decreased the lagged coherence connectivity in the delta frequency band between the right insula and left anterior cingulate cortex (ACC) (t=-3.542, p=.048). The l-DLPFC a-tDCS compared to s-tDCS decreased the lagged coherence connectivity in the delta frequency band between the right insula and left ACC (t=-4.000, p=.017). In the EO condition, the l-DLPFC a-tDCS compared to M1 s-tDCS increased the lagged coherence connectivity between the l-DLPFC and left ACC in the theta band (t=-4.059, p=.048). Regression analysis demonstrated that the a-tDCS effect on the l-DLPFC was positively correlated with sleep quality, while a-tDCS on l-DLPFC and M1 s-tDCS were positively correlated with pain catastrophizing. The application of a-tDCS over the l-DLPFC has modulated the connectivity between various brain regions involved in the affective-attentional aspects of pain, especially at lower EEG frequencies during the resting state. These findings suggest that the effects of a-tDCS on neural oscillations could serve as a neural marker associated with its impact on fibromyalgia symptoms.

## Introduction

Fibromyalgia (FM) is a primary chronic pain condition classified as nociplastic pain due to the absence of a straightforward pathophysiological process and intense emotional distress [1]. Its symptoms include musculoskeletal pain, fatigue, non-restorative sleep, cognitive changes, depressive symptoms, and other correlates of autonomic dysfunction, such as irritable bowel syndrome and bladder tenesmus [2]. The exact mechanism underlying this syndrome is not fully understood, but increased facilitatory modulation and dysfunctional descending inhibitory pathway activity may contribute to central sensitization [3]. Neuroimaging studies provide further insight into the underlying central nervous system (CNS) changes in central sensitization, demonstrating significant structural, chemical, and functional alterations in pain-processing regions such as the thalamus, periaqueductal gray (PAG), insula, cingulate, and somatosensory cortices [4, 5].

The neural network underlying the pain experience emphasizes the dynamics of functional connectivity (FC) that underpin the pain experience [6]. FC is the degree to which activity between a pair of brain regions covaries or correlates over time. In FM, the descending pain modulatory system (DPMS) does not work as well when there is more functional connectivity (FC) between pain processing areas like the left motor cortex (MC) and the left prefrontal cortex (PFC), as well as between the left MC and the right PFC [7]. Another study in FM found an increased delta value of FC in response to acute pain between the PFC and MC in genotypes BDNF Val/Met compared to Val/Val [8]. In contrast, the Val/Val group showed decreased interhemispheric connectivity in these areas, less efficiency of inhibitory DPMS, and a higher impact of fibromyalgia symptoms on quality of life [8]. An analysis of frequency maps using electroencephalography (EEG) in FM revealed anomalous FC at the bilateral precuneus, with right predominance, on the right inferior parietal cortex, bilateral prefrontal cortex, medial cortex, and right anterior cingulate cortex (ACC) [9]. Coherence analysis of the brain signal showed apparent differences between FM and controls, particularly in the bilateral frontotemporal region. Discriminatory analysis indicated a significant difference in group interconnectivity patterns [9].

Even though there is evidence that dysfunctional processing in FM is caused by dysfunctional connectivity and dysfunction of the DPMS [7,10], there is not much evidence about how treatment affects these neural markers. Thus, it is crucial to investigate the effect of treatments on these neural markers associated with the severity of pain symptoms, which can help develop effective treatments for pain management and personalized treatment. Such neural features, including cerebral oscillations and connectivity in response to treatment, can provide insights into the mechanisms of action of the treatment and whether its efficacy in clinical outcomes is related to changes in these neural markers. This way, personalized therapies based on each patient’s neuroplasticity state can be tailored. Following this rationale, it is reasonable to investigate the effect of neuromodulatory therapies that counter-regulate dysfunctional neuroplasticity underpinning chronic pain. Among these therapies is transcranial direct current stimulation (tDCS), which modulates neural cortical and subcortical networks. According to prior studies in FM, long-term tDCS at home (sixty sessions) effectively reduced analgesic use by 55% and reduced pain scores [11]. Along the same line, other trials with FM found that four weeks of home-based tDCS (HB-tDCS) improved pain-related catastrophizing [12] and cognitive impairment [13]. This top-down effect of tDCS on clinical outcomes might be related to the target area stimulated and the current type (anodal or cathodal).

The primary motor cortex (M1) modulates neural networks associated with the somatosensory system through cortical effects in the thalamic nucleus, anterior cingulate cortex (ACC), and brainstem [14]. Anodal tDCS of M1 induces corticothalamic inhibition of the ventral posterolateral nucleus (VPL), which is responsible for discriminatory sensitivity, and the ventral posteromedial nucleus (VPM), which is responsible for nociceptive sensation. Stimulation of the dorsolateral prefrontal cortex (DLPFC) decreases the activity of the midbrain-medial thalamic pathway, which is involved in the modulation of structures related to the emotional perception of pain [14]. Thus, the effect of tDCS depends on the site to be stimulated (DLPFC or M1) and the type of stimulation (anodic or cathodic) and is likely related to the number of stimulation sessions. Even though there is growing evidence of the clinical benefits of tDCS, there needs to be more literature regarding its neurophysiological effects on areas involved in pain processing and oscillatory brain signatures. Thus, additional studies are required to better understand the mechanisms of tDCS as a treatment for pain and correlated symptoms.

This study aimed to test the hypothesis that self-administered active HB-tDCS, used for 20 sessions over four weeks, would improve brain oscillations associated with resting-state EEG more than sham-(s)-tDCS. Specifically, the study investigated the effects of anodal-(a)-tDCS on the left DLPFC and a-tDCS on the primary motor cortex (M1). Additionally, we examined how lagged coherence connectivity may be related to the severity of clinical symptoms and brain-derived neurotrophic factor (BDNF) levels. We also assessed protocol adherence through valid session registry software. We hypothesized that active treatments would be superior to their respective shams in improving cerebral oscillations associated with the pain networks in areas involved in pain processing and clinical measures.

## Materials and methods

### Study design and eligibility

The trial’s protocol for the Certificate of Presentation for Ethical Appreciation (CAAE) registry number is 36995020.3.0000.5327, and the study was approved by the Research Ethics Committee at the Hospital de Clínicas de Porto Alegre (HCPA), Brazil, under registration in the Institutional Review Board (IRB) number 2020-0369. Data collected were achieved in the outpatient departments of the clinical research center of HCPA and in the research rooms of the laboratory of pain and neuromodulation of HCPA. Informed consent was obtained from all participating patients, who provided verbal and written consent to participate in the trial. The study followed a randomized, double-blind, and sham-controlled design, ensuring that neither the patients nor the researchers involved knew the treatment assignment. It is important to note that the participants in the trial did not receive any form of compensation for their involvement.

The study started in September 2019 but was paused in March 2020 due to the COVID-19 pandemic. However, in November 2020, the study resumed with several modifications and restrictions to prioritize the safety of participants and researchers. Data collection was completed in November 2022. The collected data were available on the public repository figshare DOI: https://doi.org/10.6084/m9.figshare.23542116.v1.

We adjusted the study procedures to address the challenges posed by the pandemic. Specific hospital visit dates were scheduled, and the duration of visits for psychophysical measures and device-related activities was limited. Initially, the protocol required formal consent and face-to-face assessments to gather information on clinical symptoms and demographic characteristics. However, in response to the pandemic, online methods were implemented for information collection, prioritizing the health and well-being of all involved in the study. Participants who had contracted the virus before the data collection phase was temporarily paused were placed on hold until they received a negative COVID-19 test result.

Once they tested negative, they underwent an evaluation to determine their eligibility for inclusion in the study. These adaptations were implemented to ensure the research could continue without interruption while adhering to the necessary safety measures during the pandemic.

### Inclusion and exclusion criteria

We included in the study women aged 30 to 65, literate, and diagnosed with FM according to the American College of Rheumatology (ACR) 2016 [15]. Participants needed to report a score of 6 or higher on the Numerical Pain Scale (NPS 0-10) most of the time in the last three months. Subjects were excluded in cases of pregnancy, if they had a neurological disease, a history of head trauma or neurosurgery, or a history of alcohol or drug abuse in the last six months. Besides, participants could not have decompensated systemic diseases, chronic inflammatory diseases, uncompensated hypothyroidism, another metabolic disease, or be getting treatment for cancer.

We screened 133 FM participants eligible to participate in this study. However, 85 were excluded for different reasons, such as living far away from the research center, having trouble getting around on public transportation, being unemployed, etc. Some of the screened participants did not fulfill the diagnostic criteria for FM. Besides, they were excluded if their pain levels were lower than 6 (NPS 0–10) or they had another uncompensated clinical disease (rheumatoid arthritis, lupus, hypothyroidism, etc.). So, 48 FM were included in the study, but 5 of them were excluded. So, at the end, 43 subjects were included in the analysis. The sequence of screening and assessments is presented in Fig 1.

**Fig 1.**
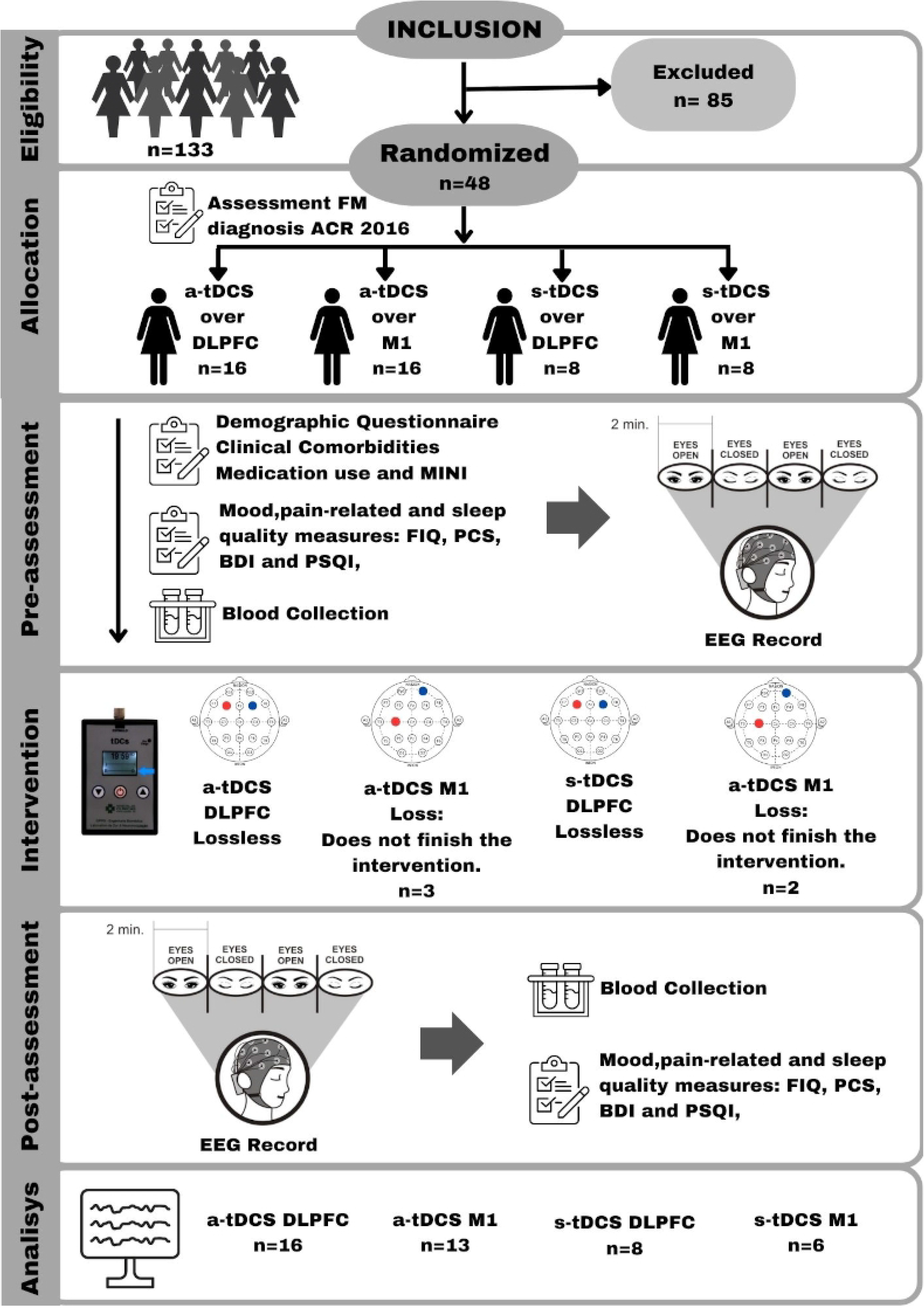
Flowchart of the study assessments. FM = Fibromyalgia; ACR = American College of Rheumatology; FIQ = Fibromyalgia Impact Questionnaire; BDI = Beck Depression Inventory; MINI = Mini International Neuropsychiatric Interview; BP-PCS = Pain Catastrophizing Scale; BP-CSI = Central sensitization inventory—Brazilian Portuguese version; PSQI = Pittsburgh Sleep Quality Index; BDNF = brain-derived neurotrophic factor; EEG= Electroencephalography; a-tDCS = active transcranial direct current stimulation; s-tDCS= sham transcranial direct current stimulation; DLPFC = Dorsolateral prefrontal cortex; M1= Motor cortex.

### Sample size justification

The sample size was estimated based on the effect seen in a previous study [16] that looked at how one session of transcranial direct current stimulation (tDCS) on the DLPFC affected EEG resting-state measurements. The study reported an effect size (η^2^ = 0.10) in the theta frequency band in the active tDCS group compared to the sham group. An analysis of variance (ANOVA) with a one-way repeated measures design was planned to determine the sample size for the current study, considering both within-group and between-group effects. The effect size for the ANOVA was set at f = 0.33, corresponding to the effect size (η^2^ = 0.10). The significance level (α) was established at 0.05, with an error (β) equal to 0.90. Based on these parameters, the sample size was determined to be 40 patients. We added 20% to account for potential dropouts, resulting in a final sample size of 48 patients. These patients were allocated into four unequal groups, with 16 patients in the active tDCS groups targeting the M1 and DLPFC regions and eight in the s-tDCS groups targeting the M1 and DLPFC regions.

### Randomization

To allocate patients to receive either a-tDCS or s-tDCS over the M1 or the DLPFC, a randomization process was conducted using the website www.sealedenvelope.com. The allocation ratio was set at 2:1:2:1, with 16 patients assigned to receive a-tDCS (either over M1 or DLPFC) and eight patients assigned to receive s-tDCS (either over M1 or DLPFC). Randomization was performed in six blocks of eight to ensure unpredictability and maintain blinding during the allocation process. Two investigators, who were not involved in the patient’s assessments, conducted the randomization process before the recruitment phase. The randomized numbers were placed inside sealed, brown envelopes, with the patient’s entry sequence number written outside each envelope. The engineer was responsible for programming the intervention device and opened the sealed envelope corresponding to the patient after obtaining informed consent. This process ensures that the allocation remains concealed until the appropriate time.

### Blinding

All participants were unaware of which stimulation type (active or sham) they received during the entire study sequence. The s-tDCS group’s device was set up to provide the stimulation three times: in the initial 30 seconds, in the middle 30 seconds, and in the last 30 seconds of the stimulation. After the post-assessment, we evaluate the blinding effectiveness by asking all participants which stimulation they suppose they received, active or sham, and measuring their confidence level using a 5-point Likert scale.

### Intervention

Participants received the programmed home-based transcranial direct current stimulation device (HB-tDCS). The treatment was performed during weekdays over 4 weeks, totaling 20 sessions. Three devices were misprogrammed and exceeded the pre-defined number of sessions. We decided not to withdraw these data from the analysis because they did not represent significant differences in the number of sessions per group. The current applied was 2 mA for 20 minutes to the active stimulation delivered using the anode electrode positioned over the left primary motor cortex (M1) and cathodal stimulation over the contra-lateral supra-orbital area for the first assembly. In the second montage, anodal stimulation was placed over the left (l)-DLPFC) and cathodal stimulation over the right DLPFC. The active a-tDCS condition is composed of a ramp-up time of 30 s for the current to go from zero to 2 mA and a ramp-down time that also takes 30 s for the current to go from 2 mA to zero to end the stimulation. For sham s-tDCS conditions, the montage was the same as a-tDCS. A 30-s ramp-up in intensity from zero to 2 mA was used for a-tDCS and s-tDCS stimulation, as well as a ramp-down for about the same duration, as explained in the blinding session. To improve the current supplied was used two silicone cannulas were attached to 35 cm^2^ (5×7 cm) electrodes coated in sponges wet with saline solution. The home-based tDCS safety was evaluated using a questionnaire based on previously reported adverse events. Details about the protocol can be seen in S2_Check list to use tDCS at home protocol and in the HB-tDCS updated protocol [17].

### Outcomes and Instruments of assessments

The primary outcome was the lagged coherence connectivity in different EEG frequency bands (delta, theta, alpha-1, alpha-2, beta-1, beta-2, beta-3, and gamma) at resting state. These measurements were taken between regions of interest (ROIs) in the pain network. The assessments were conducted in a specific sequence, as shown in Fig 1.

### Assessment of primary outcome

The electroencephalography was recorded using 18 scalp sites according to the 10–20 system, FP1, FP2, F7, F3, Fz, F4, F8, T7, C3, Cz, C4, T8, P7, P3, Pz, P4, P8, Oz, and the left ear (EXT), with reference to the right ear (CMS/DRL). The procedures were conducted with subjects sitting in a comfortable armchair, in a quiet place, and with the light of the room off. The EEG system used was the ENOBIO 20, Neuroelectrics (Barcelona, SP), which comprises an EEG cap with circular gel electrodes with a contact area of 1.75 cm^2^. Impedance was 5 kΩ for all electrodes, with high dynamic resolution (24 bits, 0.05uV) and a sampling rate of 500 Hz. A line noise filter (60-Hz) was applied to remove main line artifacts from the EEG data.

We used the EEG resting-state paradigm collected for 8 minutes total, with 2 minutes switched between EC and EO conditions. Participants were instructed to remain awake, relaxed, and thinking-free. During the EO condition, participants were instructed to fixate their vision on a small cross presented in front of them. The EC condition is the level of arousal at rest, and the EO condition is the level of arousal at activation [18,19].

### Preprocessing and functional connectivity analysis

The EEG data preprocessing was conducted with the open-source toolbox EEGLAB 14.1 [21], which ran in the MATLAB environment (The MathWorks Inc., Natick, Massachusetts, United States). Artifact detection was performed through visual inspection, removing segments of bad channels, if necessary. Continuous EEG data was band-pass filtered using a simple FIR filter with cutoff frequencies of 0.5–40 Hz, resampled to 250 Hz, and split into 4.096 s epochs.

Epochs containing artifacts were automatically excluded from the analysis. Rejection thresholds were fixed according to artifact characteristics from non-filtered continuous EEG as following [21]:

a. 50µV thresholds were set for FP1 and FP2 electrodes and 100µV thresholds for other electrodes to eliminate eye blinks and other quick movements from non-filtered continuous EEG were eliminated,
b. 50µV thresholds were used for slow waves (0-1 Hz band), while 30µV thresholds were used for fast waves (20-35 Hz band) to eliminate artifacts associated with slow head or body movements,

Brain connectivity was computed establishing the minimum threshold to be 40 seconds for each resting-state condition (EO, EC) [22]. Subjects or conditions below this threshold was rejected for the analysis.

Standardized low-resolution brain electromagnetic tomography (sLORETA) was used to measure the functional connectivity, which compute the linear dependence (coherence) of the electric neuronal activity from several brain regions [23, 24, 25]. The LORETA-Key software is freely available (https://www.uzh.ch/keyinst/NewLORETA/Software/Software.htm).

EEG electrodes coordinates employed by the software are based on the MRI anatomical template from Montreal Neurological Institute (MNI152) which slice and classify the neocortical volume (limited to the gray matter) in 6239 voxels of dimension 5mm^3^ [26, 27, 28].

However, due to the volume conduction and the low spatial resolution of EEG, the dependence measures are highly contaminated with instantaneous, non-physiological effects. In this sense, was developed of capable coherence measures that appraise the lagged connectivity between various brain locations to verify the existence of distributed cortical networks. Lagged coherence connectivity expresses the coherence measured by the corrected standardized covariance of scalp electric potentials, extracting the instantaneous linear dependence [25, 29]. The lagged coherence becomes the adequate way to measure electrophysiological connectivity, removing the confounding effect of instantaneous dependence due to volume conduction and low spatial resolution [25].

The sLORETA functional images of lagged coherence were computed to follow the frequencies band: delta (1±3.5 Hz), theta (4±7.5 Hz), alpha-1 (8±10 Hz), alpha-2 (10-12Hz), beta-1 (13±18 Hz), beta-2 (18.5±21 Hz), beta-3 (21.5±30 Hz), and gamma (30.5±44 Hz).

To generate the regions of interest (ROIs) a voxel-wise approach was implemented, MNI coordinates of the areas under the electrode were determined by sLORETA. A size 10-mm-diameter sphere was defined from the seed points centered in the following ROIs [30, 31]: left and right primary somatosensory cortex (BA01) [32], the left and right insular cortex (BA47-BA48) [33, 34], the left and right anterior cingulate cortex (BA24) [35], and the left and right dorsolateral prefrontal cortex (BA09-BA10-BA46) [35], according to previous studies on the pain network. The coordinates are shown in Table 1.

**Table 1.** Montreal Neurological Institute (MNI) coordinates of regions of interest (ROI).

| Seed | MNI coordinates |  |  | Brodmann Area |
| --- | --- | --- | --- | --- |
|  | x | y | z |  |
| Left S1 | -48 | -24 | 52 | BA01 |
| Right S1 | 52 | -16 | 44 |  |
| Left INS | -48 | 12 | -2 | BA47 and BA48 |
| Right INS | 36 | 6 | 6 |  |
| Left ACC | -2 | 8 | 30 | BA24 |
| Right ACC | 1 | 8 | 30 |  |
| Left DLPFC | -2 | 46 | -16 | BA09, BA10 and BA46<sup>d</sup> |
| Right DLPFC | 2 | 46 | -16 |  |

ACC stands for anterior cingulate cortex, INS for insula, DLPFC for dorsolateral prefrontal cortex, and S1 for primary somatosensory.

The artifact-free EEG intervals were converted into ASCII files and then put into the sLORETA software. The segmented EEG intervals recorded in resting-state were analyzed by means of a fast Fourier transform algorithm, with 2 seconds of intervals according to the following frequencies: delta (2±3.5 Hz), theta (4±7.5 Hz), alpha-1 (810 Hz), alpha-2 (10-12Hz), beta-1 (13±18 Hz), beta-2 (18.5±21 Hz), beta-3 (21.5±30 Hz), gamma (30.5±44 Hz) [23].

### Pain measures and correlated symptoms, sleep quality, depressive symptoms, socio-demographic variables, clinical and psychiatric disorders, central sensitization, and serum BDNF

All instruments utilized to evaluate pain and correlated measures, psychological tests, and sleep quality are validated for the Brazilian population, including the Fibromyalgia Impact Questionnaire (FIQ) [36], Beck Depression Inventory-II [37], the Brazilian Pain Catastrophizing Scale (BP-PCS) [38], and the Pittsburgh Sleep Quality Index (PSQI) [39].

a. The Fibromyalgia Impact Questionnaire (FIQ) was used to assess the impact of fibromyalgia on the quality of life. It includes questions about how symptoms interfere with work, including housework, as well as aspects like memory, anxiety, balance, and sensitivity to various stimuli. The maximum total score on the FIQ is 100 [36].
b. To evaluate global pain experienced over a 24-hour period throughout the four weeks of treatment, a Numeric Pain Scale (NPS) was utilized. The NPS measures pain intensity on a scale from 0 to 10, where 0 represents no pain and 10 represents the worst pain experienced in the majority of the 24-hour period across most days of the week.
c. The Pittsburgh Sleep Quality Index (PSQI) evaluates sleep quality and disturbances over the past month. The sum of the scores from seven components yields the global PSQI score, which ranges from 0 to 21. A higher score indicates poorer sleep quality [39].
d. The Beck Depression Inventory - Second Edition (BDI-II) is a self-report instrument consisting of 21 items. Its purpose is to measure the intensity of depression [37].
e. The Brazilian Portuguese translation of the Pain Catastrophizing Scale (BP-PCS) comprises 13 items divided into three domains: magnification, helplessness, and rumination. The questions aim to assess the patient’s feelings and thoughts related to pain and measure pain-related catastrophizing [38].
g. Enzyme-linked immunosorbent assay (ELISA) was used to measure blood levels of BDNF. Monoclonal antibodies specific for BDNF were employed from R&D Systems (MN, USA; ChemiKine BDNF Sandwich ELISA kit, CYT306; Chemicon/Millipore, Billerica, MA, USA). The inter-assay variance was evaluated using two plates per kit on two different days within the same week. All procedures followed the manufacturer’s recommendations, and the lowest detection limit for BDNF was 7.8 pg/ml. Optical density measurements at 450 nm were conducted using the Promega GloMax®-Multi Microplate Reader. Additionally, the Bio-Plex®-200 instrument from Bio-Rad was used for multiplexing assay measurements. Total protein was assessed using bovine serum albumin following the Bradford method.
h. A standardized questionnaire was utilized to collect demographic data and medical comorbidities. Patients were asked to furnish details concerning their age, gender, educational background, marital status, and lifestyle choices. Additionally, they provided information about their overall health condition, including clinical and psychiatric diagnoses.
i. Mini-International Neuropsychiatric Interview (MINI) is a short (15–30 min) structured psychiatric diagnostic interview aimed to screen for DSM-IV and ICD-10 diagnoses [40].
j. The central sensitization inventory (CSI) is a tool that identifies key symptoms related to central sensitization processes by quantifying the severity of these symptoms. It consists of two parts: Part A is a 25-item self-report questionnaire designed to assess symptoms related to health and Part B (not rated) is designed to determine if one or more specific disorders [41].

### Statistical Analysis

Descriptive statistics were employed to summarize the main demographic characteristics of the sample. The Shapiro-Wilk test was used to assess the data distribution. To compare continuous variables between groups, independent sample t-tests were utilized. The chi-squared and Fisher’s exact tests were employed for comparing categorical variables between groups. Within-group analyses were conducted using paired t-tests to examine differences between baseline and treatment end measures.

Significant differences in lagged coherence for each frequency band across the eight regions of interest (ROIs) were determined using a t-test for multiple comparisons. These differences were utilized to construct a correlation matrix for generating connectivity maps. Statistical nonparametric mapping (SnPM) with 5000 permutations was performed to identify cortical voxels that exhibited significant differences.

The correlation matrices were used to see how stimulation affected changes in lagged coherence when the eyes were EO or EC at different levels of analysis. We compared lagged coherence within groups in the EO and EC conditions.

Linear regressions were performed separately for the eyes open (EO) and eyes closed (EC) conditions. A test with 5000 permutations was utilized to determine the significance threshold and correct for multiple comparisons. All analyses were conducted using two-tailed tests, and a type I error of 5% was accepted. A two-tailed type I error of 0.05 was used for all analyses. The statistical analysis was conducted using the sLORETA software. All statistical analyses were performed using two-tailed tests at the 5% significance level with SPSS, version 22.0 (SPSS, Chicago, IL).

## Results

### Demographic and clinical characteristics of the subjects

We included 48 women with fibromyalgia. However, we examined a total of 43 participants who were randomly assigned to one of the following treatment groups: a-tDCS on DLPFC (n=16), a-tDCS on M1 (n=13), s-tDCS on DLPFC (n=8), or s-tDCS on M1 (n=6). Five patients were excluded from the analyzes. Table 2 provides an overview of the subjects’ baseline demographic and clinical characteristics.

**Table 2.**
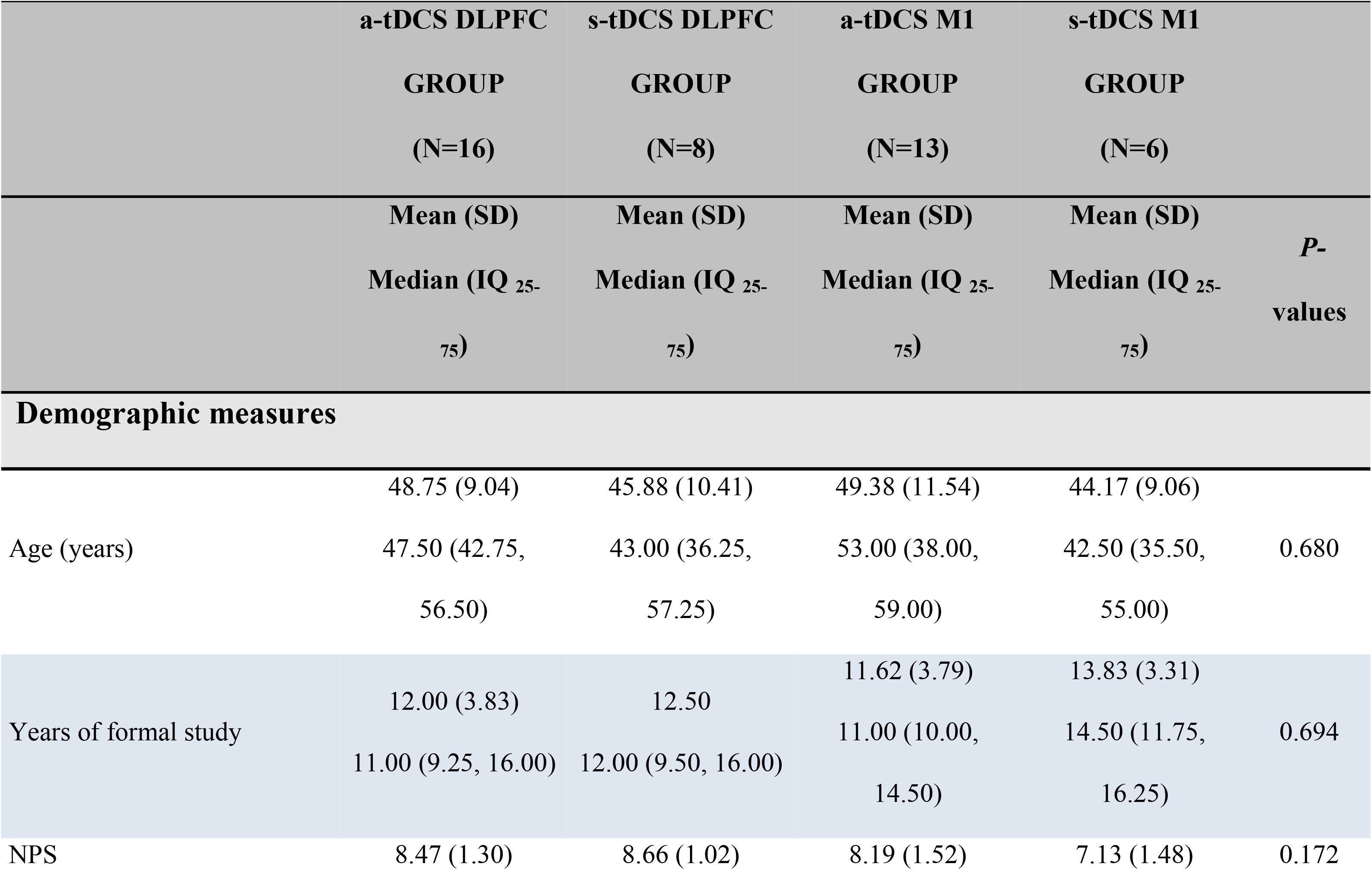
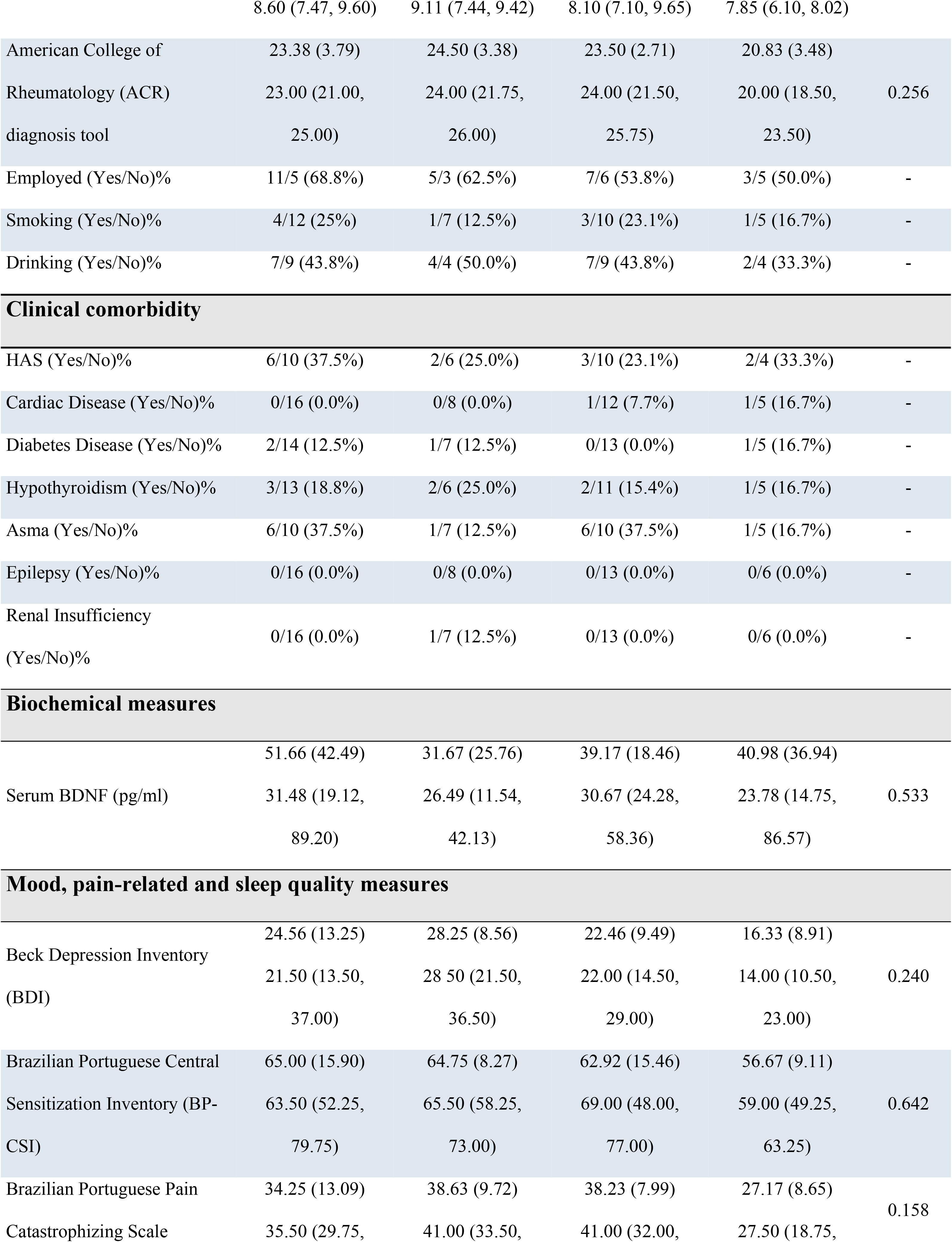
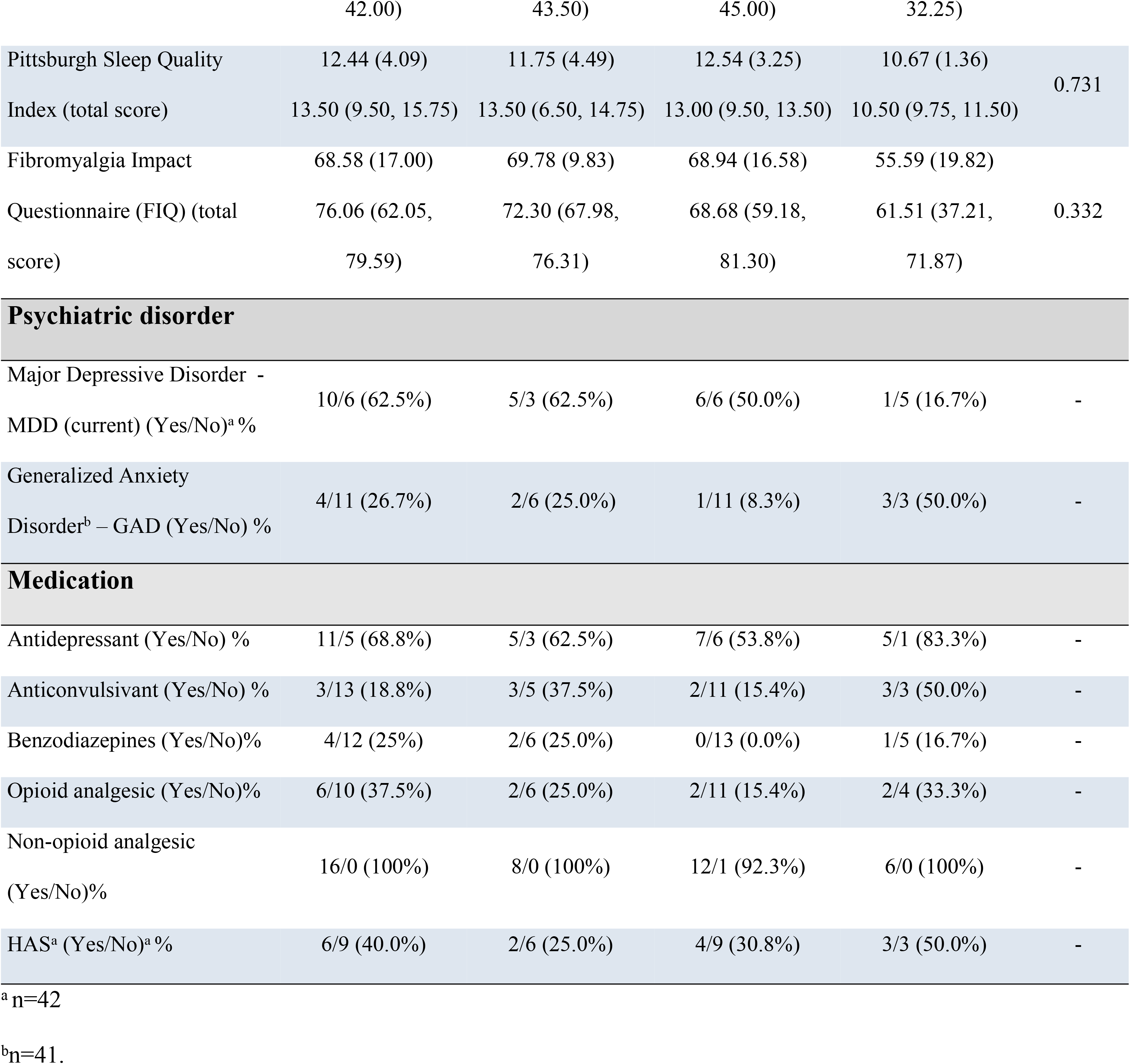
Demographic and clinical characteristics of the study sample are presented. Values are reported as mean (SD) for continuous variables, median (IQ) or % for categorical variables (n=43).

## Primary outcome

### Effect of treatment between-groups on lagged coherence connectivity in EC and EO conditions

In the EC condition, when it was assessed the comparisons of lagged coherence connectivity expressed by Δ-means (pre-minus post-treatment) according to the treatment group (a-tDCS and s-tDCS) applied over left DLPC, we found that the a-tDCS over left DLPFC decreased connectivity between right insula and left ACC in the delta frequency band (see Fig 2A). In EC and EO conditions, statistical significance was not identified between a-tDCS vs. s-tDCS over DLPFC or M1 in all frequency bands.

**Fig 2.**
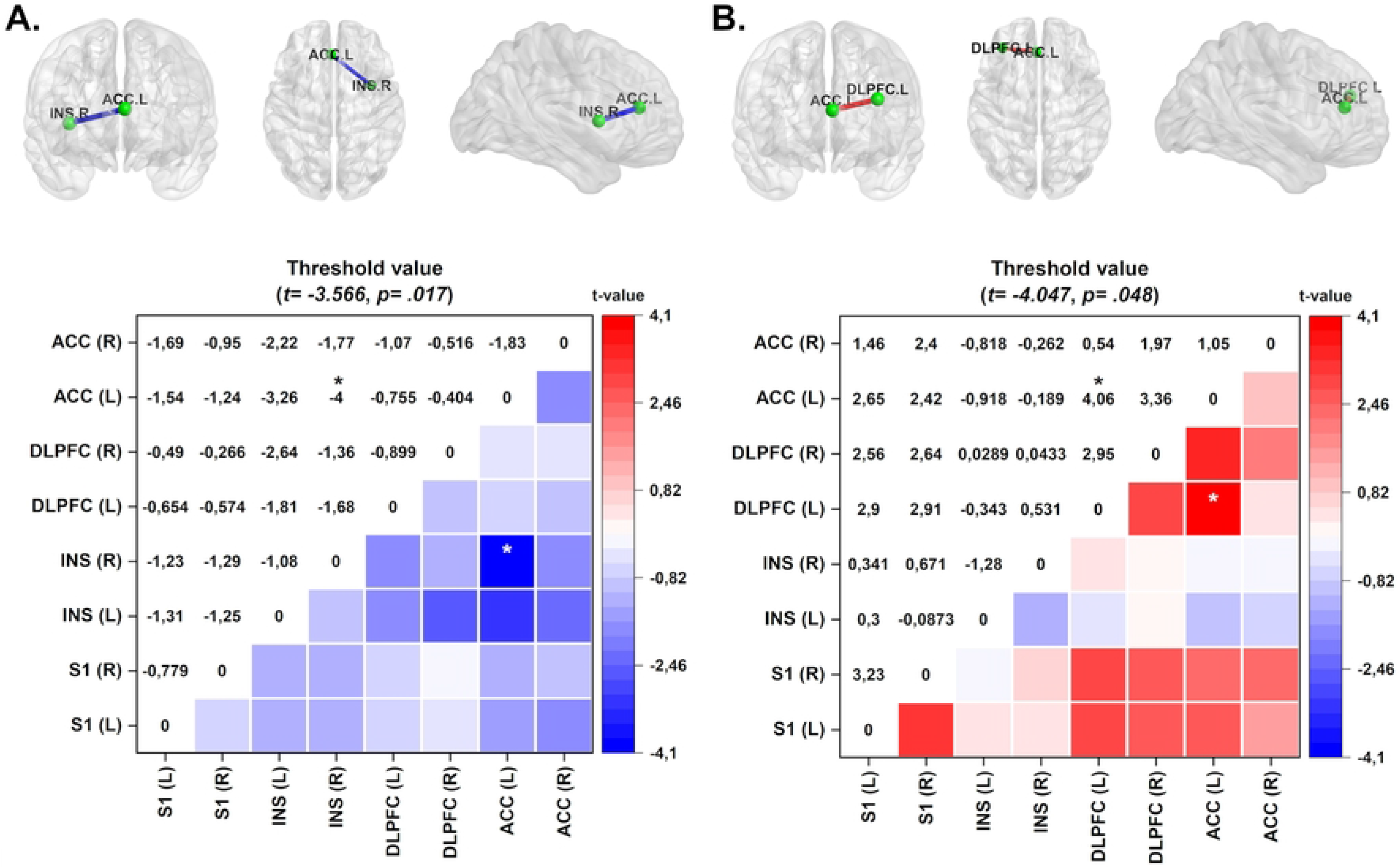
Connectivity maps and color maps of between-group analysis of the tDCS intervention. The colored edge represents connections with significant differences. (A) Maps shows decreased connectivity between left ACC and right INS in the a-tDCS on DLPFC group in the delta frequency band. (B) a-tDCS over DLPFC group exhibits decreased connectivity in the theta frequency band between the left DLPFC and left ACC. Values are given in t-values. ACC = anterior cingulate cortex; INS = insula; DLPFC = dorsolateral prefrontal cortex; SI = primary somatosensory cortex. *p<0.05.

In the EO condition, when it was assessed, the comparison of lagged coherence connectivity expressed by Δ-means (pre-minus post-treatment) according to the treatment group a-tDCS applied over the left DLPFC exhibits increased connectivity in the theta frequency band between the left DLPFC and left ACC compared with sham-tDCS (pre-minus post-treatment) over M1 (see Fig 2B). In contrast, in the EC condition, we did not find a difference between these groups.

The lagged coherence analysis in EO and EC conditions doesn’t show differences in the a-tDCS pre- to post-treatment (Δs) between DLPFC compared to M1, as well as between a-tDCS over M1 compared with s-tDCS over DLPFC.

### Effect of treatment within-groups on lagged coherence connectivity in EC and EO conditions

In the EC condition, when it was assessed the comparisons of lagged coherence connectivity expressed by Δ-means (pre-minus post-treatment) according to the treatment group (a-tDCS and s-tDCS) applied over target brain areas nominally left DLPC and M1, we found that the a-tDCS over left DLPFC decreased connectivity between right insula and left ACC in the delta frequency band (see Fig 3A). In EC and EO conditions, statistical significance was not identified in the sham DLPFC stimulation, and in active stimulation over M1, for all frequency bands.

**Fig 3.**
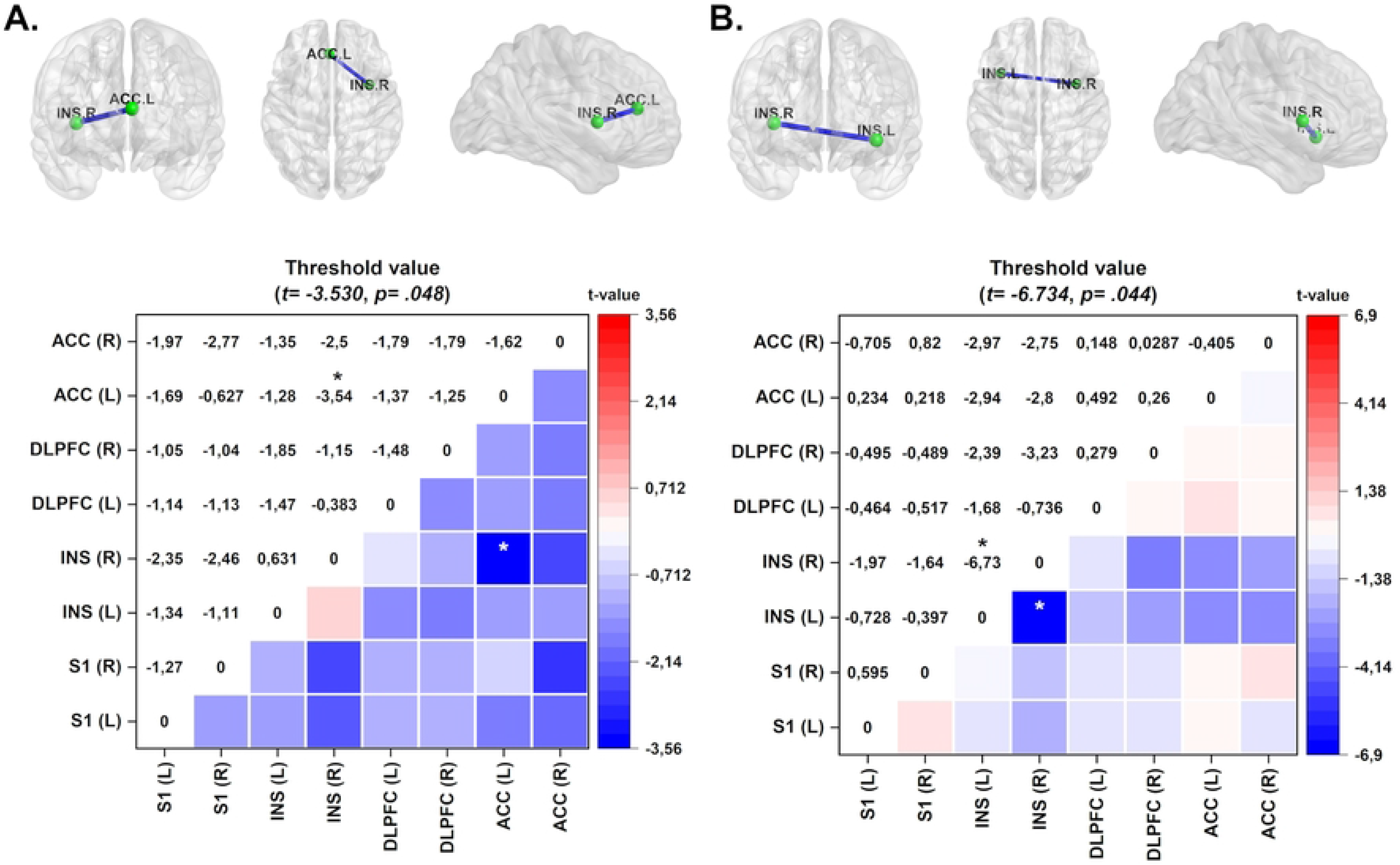
Connectivity maps and color maps of within-group analysis of the tDCS intervention. The colored edge represents connections with significant differences. Data represent the delta-value of the mean oscillation of the frequency band from pre- to post-treatment in each brain region. (A) The maps demonstrate decreased connectivity between the left ACC and right INS in the a-tDCS on DLPFC group in the delta frequency band in EO condition. (B) The s-tDCS on M1 group exhibits increased connectivity in the gamma frequency band between the right and left INS. Values are given in t-values. ACC = anterior cingulate cortex; INS = insula; DLPFC = dorsolateral prefrontal cortex; SI = primary somatosensory cortex. *p<0.05.

In the EO condition, when it was assessed the comparisons of lagged coherence connectivity expressed by Δ-means (pre-minus post-treatment) s-tDCS over on M1 demonstrated decreased lagged coherence connectivity between left and right insula in the gamma frequency band in EO condition (see Fig 3B).

## Secondary outcomes – exploratory analysis

### Effect of treatment within-groups on the delta-value lagged coherence connectivity in the EC and EO Conditions, sleep quality and pain catastrophizing

We conducted a series of linear regression analyses to investigate the relationship between ROI connectivity in specific frequency bands (delta, theta alpha, beta, and gamma) and various factors such as depressive symptoms, pain-related measures, sleep quality, and serum BDNF levels. We present the variables that showed a statistically significant correlation with the frequency bands, considering either EO or EC conditions.

In the EO condition, where participants received a-tDCS over the DLPFC, we observed a positive correlation between sleep quality and the lagged coherence connectivity in the beta-3 frequency band. Specifically, this correlation was observed between the left insula and the left and right primary somatosensory cortex (S1) (see Fig 4A).

**Fig 4.**
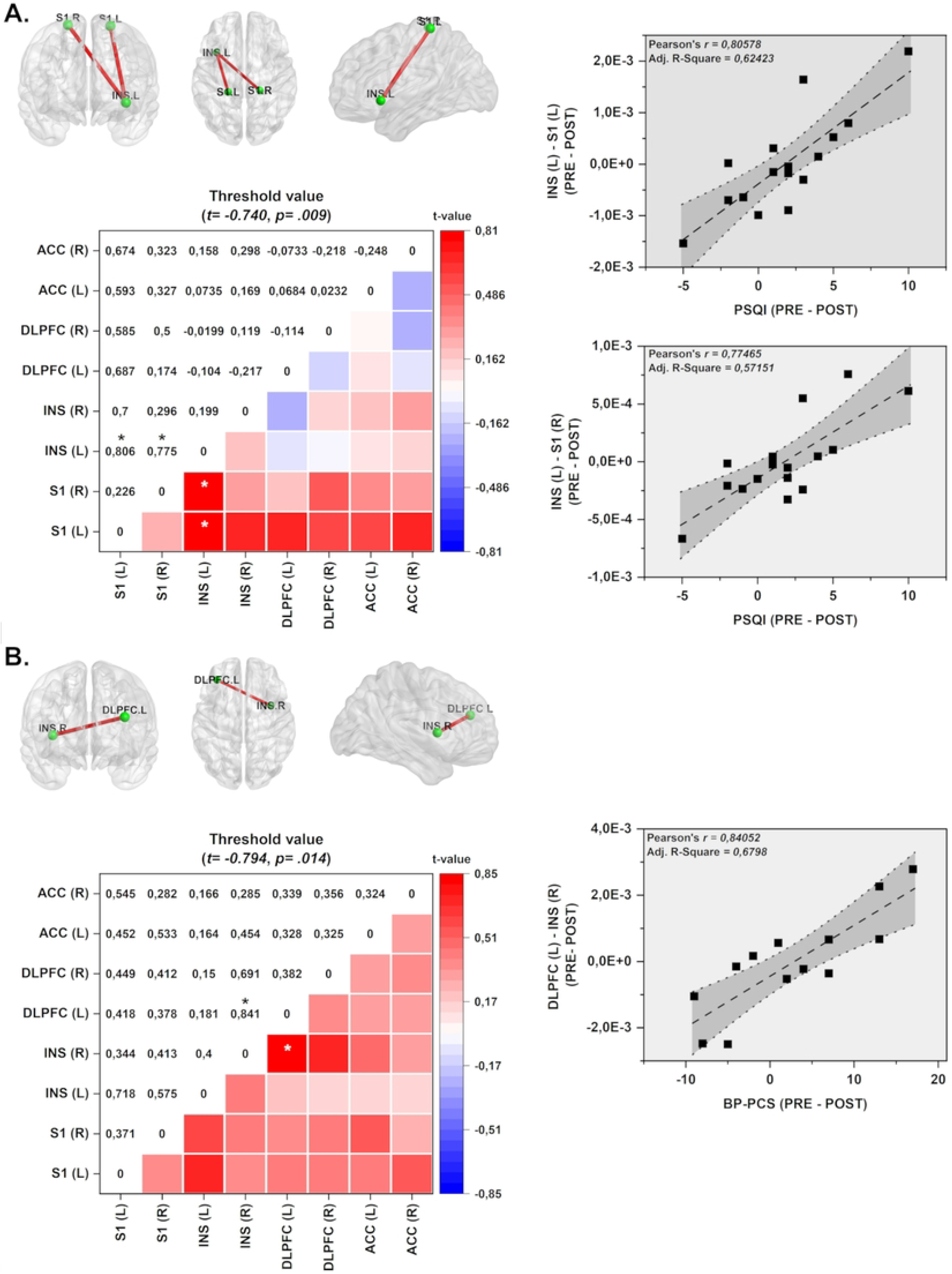
Connectivity and color maps of the linear regression analyses considering Δ-values of differences pre- to post-treatment of ROI connectivity. The Δ-values of ROI connectivity were correlated with Δ-values of the following variables: depressive symptoms, pain-related, sleep quality, and serum BDNF. (A) The a-tDCS over DLPFC group presented increased lagged coherence connectivity in the beta-3 band between left INS and left and right S1, which is correlated with sleep quality measures (PSQI) in EO condition. (B) The a-tDCS over M1 group shows increased coherence connections in the delta frequency band between right INS and left DLPFC, which is correlated with pain catastrophizing scores (Br-PCS) in the EC condition. Values given in Cohen r = correlation coefficient (small=0.1; medium=0.3; large=0.5). ACC = anterior cingulate cortex; INS = insula; mPFC = medial prefrontal cortex; S1 = primary somatosensory cortex. \**p*<0.05.

In the EC condition, where participants received a-tDCS over the M1, we found that change of pain catastrophizing form pre to treatment end was positively correlated with the lagged coherence connectivity in the delta frequency band. This correlation was observed between the right insula and the left DLPFC (see Fig 4B).

### Adherence, compliance, and side effects across the treatment

The data in Fig 5 provides information on the average number of valid sessions completed per group, indicating the level of adherence and compliance. The mean (SD) values reflect the average number of valid sessions completed, while the median (IQ_25-75_) values give the middle range of valid session counts within each group. According to the total sample size, we planned 860 tDCS sessions and recorded 758 valid sessions, corresponding to 88.1% of the total amount.

**Fig 5.**
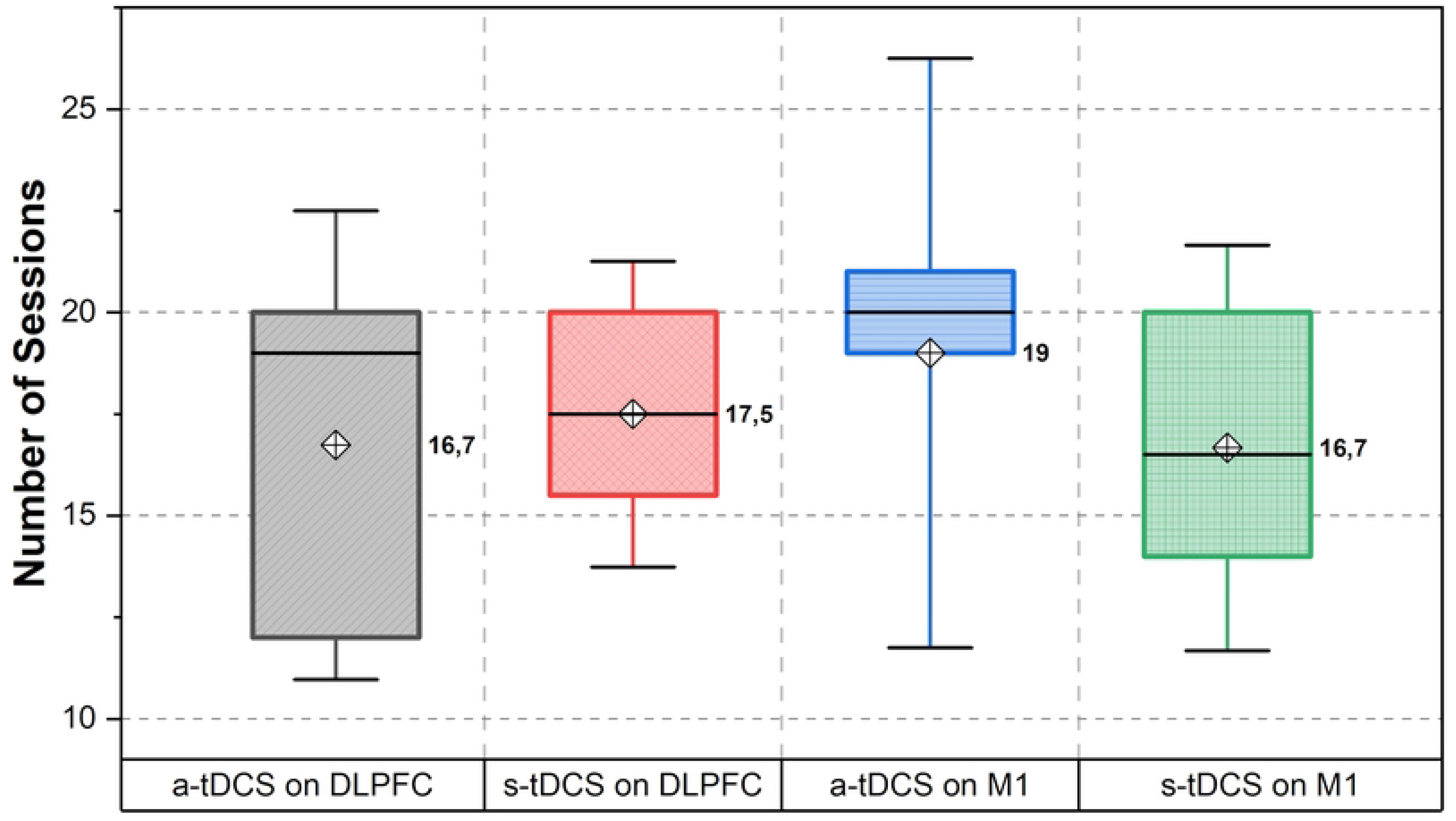
Number of sessions according to a-tDCS on DLPFC, s-tDCS on DLPFC, a-tDCS on M1, and s-tDCS on M1.

We predicted 320 sessions for the a-tDCS over the DLPFC group (n=16) and conducted 271 valid sessions that comprised 84.68% of the total. In the group where s-tDCS was applied over DLPFC (n=8), 140 of the 160 estimated sessions were completed, equivalent to 87.5%. For the group stimulated with a-tDCS on M1 (n=13), we administered 247 sessions of the 260 determined, corresponding to 95% of the total sum. Finally, of the 120 sessions stipulated by the s-tDCS on the M1 group (n=6), 100 were performed, computing 83.33%.

The side effects questionnaire included inquiries about various symptoms, including headache, neck aches, tingling, redness, burning sensation, itching, somnolence, and decreased concentration. The Fig 6 illustrates the reported symptoms and their severity classified as absence, mild moderate and severe. It is important to note that no participants discontinued the treatment due to uncomfortable side effects.

**Fig 6.**
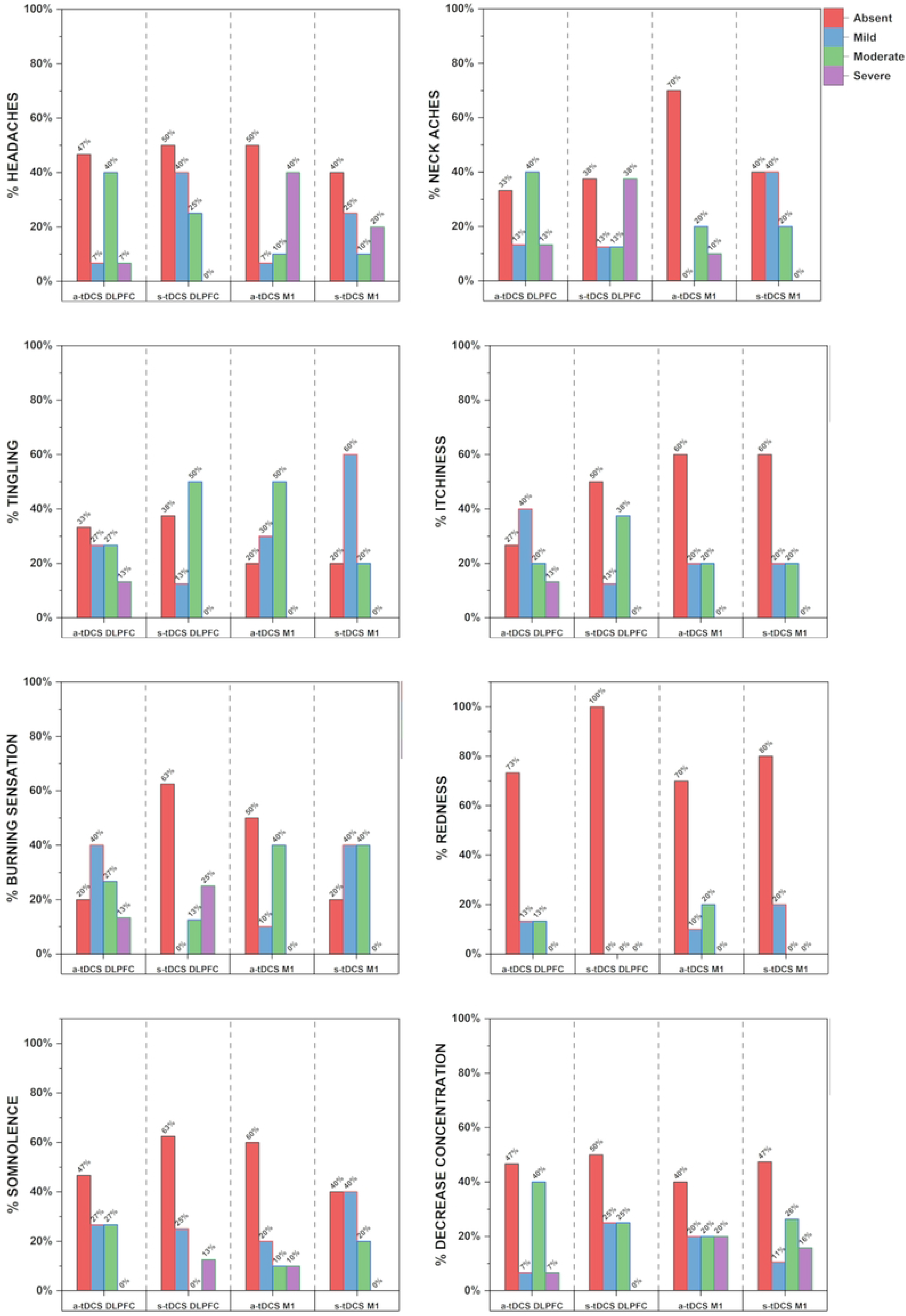
Side effects are presented as percentages (%).

## Discussion

These findings expand the evidence regarding the effects of self-applied HB-tDCS based on a trial with a factorial design in FM. The trial included twenty sessions of tDCS administered over four weeks, targeting either the left DLPFC or the M1. The originality of this study lies in its understanding of how tDCS affects frequency bands in particular brain regions which operate as neurophysiological markers of neuroplasticity and how it affects related symptoms. Besides, they showed that a-tDCS modulates different brain areas distinctly.

They revealed that a-tDCS over the DLPFC reduced connectivity within the delta frequency band between the right insula and the left ACC. These brain regions are known to be involved in cognitive and emotional processing. In pain processing, the insula encompasses sensory, affective, and cognitive aspects. According to neuroimaging studies, during the experience of pain, there is consistently heightened activity in the insula [42]. Additionally, increased glutamate levels cause the insula to become more excitable, which increases pain hypersensitivity [43]. Thus, these results provide valuable insights into the neurophysiological changes associated with the a-tDCS effects in pain processing. Remarkably, these findings align with studies using a pharmacological treatment for chronic pain, which reduced the insular cortex connectivity between these brain structures [44]. Since the insula is involved in sensory or affective aspects of pain, physiological processes also support these findings. At the same time, the ACC evaluates and modulates the emotional and cognitive aspects of pain [45]. In this way, the likely effect of a-tDCS in the interaction between these two brain regions is vital for modulating the subjective experience of pain. Since the insula is connected to other parts of the brain, such as the DLPFC, amygdala, ACC, mPFC, and orbitofrontal cortices, these findings show how a-tDCS applied over the DLPFC might change the neuroplasticity processes that underlie clinical outcomes related to pain and related symptoms.

The a-tDCS over the DLPFC group exhibited increased connectivity in the theta frequency band between the left DLPFC and left ACC under the EC condition. This result highlights that the a-tDCS over the DLPFC may enhance information integration and coordination between these two brain regions, which have been related to cognitive processes and emotional regulation [46]. So, an increased coupling (positive correlations) between these regions indicates that the improvement in the adaptive regulation of cognition and emotions is linked to the effect of treatment. This statement is supported by robust evidence of meta-analysis showing that the increased ACC activity is related to a positive antidepressant response, including various drugs (e.g., selective serotonin reuptake inhibitors (SSRIs), atypical antidepressants, ketamine), sleep deprivation, and repetitive transcranial magnetic stimulation (rTMS) [46]. The current study gives insights that highlight the a-tDCS effect on brain areas related to dysfunctional neurobiological processes of the pre-frontal cortex, particularly on the left side, which have been found in individuals with chronic pain and FM [45]. Accordingly, in earlier studies, these regions have been found to have reduced activity associated with difficulties in attentional control, increased pain perception, and emotional disturbances observed in FM [47, 48]. Although this point of discussion is inferential, these connectivity measures support the compression of the a-tDCS effect on the left PFC neural networks. Besides, the effect of a-tDCS in this target area also reduced depressive symptoms. Overall, they are aligned with specialized literature that also shows PFC dysfunction and structural changes in major depressive disorders [49]. However, the interpretation of these results requires parsimony. Further research is needed to investigate if these remaps of neural networks in pathways associated with pain processing have a relevant clinical impact on patient-reported outcome measures (PROM).

The a-tDCS on the DLPFC decreased connectivity between the left ACC and right insula in the delta frequency band under the EO condition. In contrast, the s-tDCS in the M1 group exhibited decreased connectivity in the gamma frequency band between the right and left insula under the EO condition. These findings indicate that distinct factors might modulate the connectivity between brain regions besides of effect of electric current applied during active t-DCS. The effect found on the ACC and right insula suggests that a-tDCS can modulate the affective-attentional circuits in FM. They agree with the idea from earlier studies [50, 51] that dysfunctional neuroplasticity linked to negative emotions and seen in chronic pain needs to be countered by regulating excitatory activity in the ACC neurons. So, they offer evidence to support the biological plausibility of the relationship between the a-tDCS effect and clinical symptoms related to FM, such as the improvement in cognitive impairment, pain catastrophizing, and depressive symptoms [12, 13, 52]. Previous studies that show a connection between the oscillations in the delta frequency and cognitive functions like attention, working memory, and emotional regulation support this conclusion [53]. Thus, the increased connectivity between the left DLPFC and left ACC in the theta frequency band implies enhanced communication and coordination between these regions, which are known to be involved in emotional and cognitive processing. In this way, these findings are in congruence with earlier studies that found that excitatory activity in ACC neurons is necessary for the experience of pain-related negative emotions. Hence, they are aligned with the theoretical perspective of earlier research, which suggests that modulating the ACC’s activity can impact the emotional aspects of pain perception [54]. As a result, a decrease in pain catastrophizing scores from pre-treatment at the end of treatment suggests that there may be a connection between these two brain areas and clinical improvement. This result backs up earlier studies that showed an a-tDCS effect in the left dorsal ACC improved not only pain disability but also depression in patients with chronic low back pain [55]. So, these results, which are backed up by evidence that shows both biological plausibility and the effect on clinical symptoms, could be used to support frequency band oscillations as neural markers of a-TDCS in different parts of the brain. However, future research should focus on elucidating the precise mechanisms and functional implications of these relationships in pain processing and whether the brain oscillation band might be a biomarker of treatment response.

The s-tDCS in the M1 group exhibited decreased connectivity in the gamma frequency band between the right and left insula under the EO condition. This result suggests a reduced level of synchronized activity between these two regions in EO conditions. Gamma waves are one of the fastest forms of brain activity, and several factors are related to their frequency oscillations. They can vary depending on the specific context, and we do not have a clear explanation for this finding. The balance between excitatory and inhibitory neurotransmission in the brain affects gamma oscillations. Specifically, the inhibitory neurotransmitter gamma-aminobutyric acid (GABA) is crucial in regulating gamma frequency oscillations. They are associated with various cognitive processes, including attention, working memory, sensory perception, learning different aspects of sensory information, and facilitating the formation and retrieval of memories.

We found that when the a-tDCS was put on the DLPFC, the oscillations in the beta-3 band caused the PSQI scores to drop more from the beginning of treatment to the end of treatment in the EC condition. These results revealed that the connectivity between the left insula and the right and left primary sensory cortex (S1) is positively correlated with the improvement in sleep quality score. Previous studies found that increased brain connectivity between S1 and the left insula was linked to increased self-reported attention to the pain stimulus and increased clinical pain in the chronic low back [56, 57]. Another study using fMRI found decreased resting-state functional connectivity between the left somatosensory cortex (S1) and the periaqueductal gray (PAG) (S1-PAG rs-FC). This reduced connectivity between these two structures was related to the severity of dysfunction in the descending pain modulatory system (DPMS). Also, the S1-PAG rs-FC in the left brain hemisphere was positively correlated with a central sensitization symptom and negatively correlated with sleep quality and pain scores [10]. Although we do not have a clear explanation for these findings as well as their relationship with the tDCS effect, we are aware that this is an exploratory analysis that agrees with previous research that individuals with fibromyalgia may exhibit increased connectivity patterns between different brain regions, including the insula and the S1 area [56, 57]. This relationship between reduced connectivity between the left insula and S1 and the improvement in sleep quality indicates that altered connectivity within the brain’s networks, specifically between the left insula and the DMN, may contribute to comprehending the possible impact of a-tDCS to remap the function in the neural networks, which is responsible for disrupted sleep patterns. The association between reduced connectivity and improved sleep quality suggests that it was related to the a-tDCS effect over the DLPFC.

The main concerns in interpreting these findings that must be considered are: First, it is essential to note that although patients received thorough training in using the tDCS device, we did not remotely monitor their sessions, and the researcher was available to solve any problem with the device at any time. One needs to be cautious when comparing directly with studies that employed electrode placement and exposure supervision. *Second*, the adherence rate is consistent with past research that supported this method of self-application for prolonged tDCS use at home [11, 12]. *Third*, the tDCS system utilized in this study offers an efficient technical solution that allows medical engineers, who were not directly involved in the patient’s care, to program the device according to the randomization sequence. This approach ensures the blinding of both the research team and patients, eliminating bias in the study design. *Fourth,* while the randomization procedure ensured that groups receiving active tDCS (a-tDCS) and sham tDCS (s-tDCS) were balanced, it is crucial to recognize that various confounding factors may affect electrophysiological signals. These variables include psychiatric comorbidities such as depression, anxiety, and sleep disturbances, as well as medication use, especially opioids, which can change the cortical reaction, and side effects that are difficult to entirely control [58]. *Fifth*, we have chosen to allocate participants in a ratio of 2:1 based on the rationale that fibromyalgia causes significant long-term suffering. This allocation ratio is intended to prioritize the active treatment group as we believe that by treating more individuals actively, we can potentially increase adherence to the treatment. Moreover, having a larger sample size in the active group enhances our ability to identify any potential side effects associated with the treatment [59, 60]. By implementing this allocation strategy, we aim to maximize the benefits for participants while ensuring a comprehensive evaluation of the treatment’s effectiveness and safety. *Sixth*; our study revealed a high adherence rate exceeding 85% for both active tDCS (a-tDCS) and sham tDCS (s-sham tDCS) sessions. We employed a rigorous and replicable methodology to showcase the effectiveness and practicality of tDCS in a home setting. *Seventh,* despite using a standardized protocol to collect the data, we needed to exclude five patients. This issue may reduce the power of certain analyses and increase type I and type II errors. Therefore, it is essential to interpret the outcomes of secondary variables in an explanatory manner. *Eight* studies using 18 EEG electrodes for source localization may have limitations in capturing the full complexity and fine-grained details of brain activity across different regions [61]. Despite the trade-off between the practical rules of electrode placement and spatial resolution, these data offer valuable insights into neural pain processing and correlated symptoms. Numerous studies, including those on FM, obsessive-compulsive disorder (OCD), tinnitus, and autism spectrum disorder (ASD), support this strategy [62, 30, 63–66]. Finaly, to mitigate potential bias related to sex, we specifically recruited female participants for our study. This choice was made because previous research shows that a-tDCS over the DLPFC leads to a greater flow of current toward the frontal regions in women [66], and other research shows that women tend to respond better to a-tDCS than men [67].

The application of a-tDCS over the l-DLPFC has been shown to modulate the connectivity between various brain regions involved in the affective-attentional aspects of pain, especially at lower EEG frequencies during the resting state. These findings suggest that the effects of a-tDCS on neural oscillations could serve as a neural marker that is associated with its impact on fibromyalgia symptoms.

## Data Availability

All data might be found on: https://doi.org/10.6084/m9.figshare.23542116.v1

## Author Contributions

**Conceptualization:** ILST FF WC

**Data curation:** FF WC

**Formal analysis:** RLA MZ WC

**Funding acquisition:** PRSS DPSJ ILST WC

**Investigation:** RLA PVS BFT VSL RBT RPA SMB LM

**Methodology:** RLA MZ PRSS DPSJ WC

**Project administration:** MZ LR CFSA PRSS DPSJ WC

**Resources:** PRSS DPSJ ILST WC

**Supervision:** MZ LR CFSA WC

**Visualization:** RLA WC

**Writing-original draft:** RLA MZ WC

**Writing-review & editing:** RLA WC

## Acknowledgments

The authors would like to thank all the patient volunteers and the Hospital de Clínicas de Porto Alegre for the logistic support. We also would like to thank the Laboratory of Pain and Neuromodulation staff for the effort and helps to conduct this study, mainly to great researcher Vani dos Santos. Remembering her keeps inspiring us.

## Supporting Information

**S1_File. CONSORT Checklist**

**S2_File. Check list to use tDCS at home protocol**

**S3_File. Protocol and Ethical Approval**

**S1_Table. Supporting Data**

## References

1. Treede RD, Rief W, Barke A, Aziz Q, Bennett MI, Benoliel R, et al. Chronic pain as a symptom or a disease: the IASP Classification of Chronic Pain for the International Classification of Diseases (ICD-11). Pain. 2019 Jan;160(1):19–27. doi: 10.1097/j.pain.0000000000001384. PMID: 30586067.

2. Duruturk N, Tuzun EH, Culhaoglu B. Is balance exercise training as effective as aerobic exercise training in fibromyalgia syndrome? Rheumatol Int. 2015 May;35(5):845–54. doi: 10.1007/s00296-014-3159-z. PMID: 25903448.

3. Harte SE, Harris RE, Clauw, DJ. The neurobiology of central sensitization. J Appl Behav Res. 2018; 23:e12137. https://doi.org/10.1111/jabr.12137

4. Smallwood J, Gorgolewski KJ, Golchert J, Ruby FJ, Engen H, Baird B, et al. The default modes of reading: modulation of posterior cingulate and medial prefrontal cortex connectivity associated with comprehension and task focus while reading. Front Hum Neurosci. 2013 Nov 12;7:734. doi: 10.3389/fnhum.2013.00734. PMID: 24282397.

5. Kenshalo DR Jr, Isensee O. Responses of primate SI cortical neurons to noxious stimuli. J Neurophysiol. 1983 Dec;50(6):1479–96. doi: 10.1152/jn.1983.50.6.1479. PMID: 6663338.

6. Peyron R, Fauchon C. The posterior insular-opercular cortex: An access to the brain networks of thermosensory and nociceptive processes? Neurosci Lett. 2019 May 29;702:34–39. doi: 10.1016/j.neulet.2018.11.042. Epub 2018 Nov 29. PMID: 30503920.

7. de Oliveira Franco Á, da Silveira Alves CF, Vicuña P, Bandeira J, de Aratanha MA, Torres ILS, et al. Hyper-connectivity between the left motor cortex and prefrontal cortex is associated with the severity of dysfunction of the descending pain modulatory system in fibromyalgia. PLoS One. 2022 May 27;17(5):e0247629. doi: 10.1371/journal.pone.0247629. PMID: 35622879.

8. de Oliveira Franco Á, de Oliveira Venturini G, da Silveira Alves CF, Alves RL, Vicuña P, Ramalho L, et al. Functional connectivity response to acute pain assessed by fNIRS is associated with BDNF genotype in fibromyalgia: an exploratory study. Sci Rep. 2022 Nov 6;12(1):18831. doi: 10.1038/s41598-022-23476-3. PMID: 36336706.

9. Martín-Brufau R, Gómez MN, Sanchez-Sanchez-Rojas L, Nombela C. Fibromyalgia Detection Based on EEG Connectivity Patterns. J Clin Med. 2021 Jul 25;10(15):3277. doi: 10.3390/jcm10153277. PMID: 34362061.

10. Soldatelli M, Franco ÁO, Picon F, Duarte JÁ, Scherer R, Bandeira J, et al. Primary somatosensory cortex and periaqueductal gray functional connectivity as a marker of the dysfunction of the descending pain modulatory system in fibromyalgia. Korean J Pain. 2023 Jan 1;36(1):113–127. doi: 10.3344/kjp.22225. PMID: 36581601.

11. Brietzke AP, Zortea M, Carvalho F, Sanches PRS, Silva DPJ, Torres ILDS, et al. Large Treatment Effect With Extended Home-Based Transcranial Direct Current Stimulation Over Dorsolateral Prefrontal Cortex in Fibromyalgia: A Proof of Concept Sham-Randomized Clinical Study. J Pain. 2020 Jan-Feb;21(1-2):212–224. doi: 10.1016/j.jpain.2019.06.013. PMID: 31356985.

12. Caumo W, Alves RL, Vicuña P, Alves CFDS, Ramalho L, Sanches PRS, et al. Impact of Bifrontal Home-Based Transcranial Direct Current Stimulation in Pain Catastrophizing and Disability due to Pain in Fibromyalgia: A Randomized, Double-Blind Sham-Controlled Study. J Pain. 2022 Apr;23(4):641–656. doi: 10.1016/j.jpain.2021.11.002. PMID: 34785366.

13. Serrano PV, Zortea M, Alves RL, Beltrán G, Bavaresco C, Ramalho L, et al. The effect of home-based transcranial direct current stimulation in cognitive performance in fibromyalgia: A randomized, double-blind sham-controlled trial. Front Hum Neurosci. 2022 Nov 24;16:992742. doi: 10.3389/fnhum.2022.992742. PMID: 36504629.

14. Zortea M, Ramalho L, Alves RL, Alves CFDS, Braulio G, Torres ILDS, et al. Transcranial Direct Current Stimulation to Improve the Dysfunction of Descending Pain Modulatory System Related to Opioids in Chronic Non-cancer Pain: An Integrative Review of Neurobiology and Meta-Analysis. Front Neurosci. 2019 Nov 18;13:1218. doi: 10.3389/fnins.2019.01218. PMID: 31803005.

15. Wolfe F, Clauw DJ, Fitzcharles MA, Goldenberg DL, Häuser W, Katz RL, et al. 2016 Revisions to the 2010/2011 fibromyalgia diagnostic criteria. Semin Arthritis Rheum. 2016 Dec;46(3):319–329. doi: 10.1016/j.semarthrit.2016.08.012. PMID: 27916278.

16. Kim J, Jang KI, Roh D, Kim H, Kim DH. A direct comparison of the electrophysiological effects of transcranial direct and alternating current stimulation in healthy subjects. Brain Res. 2020 Nov 15;1747:147065. doi: 10.1016/j.brainres.2020.147065. PMID: 32818525.

17. Carvalho F, Brietzke AP, Gasparin A, Dos Santos FP, Vercelino R, Ballester RF, et al. Home-Based Transcranial Direct Current Stimulation Device Development: An Updated Protocol Used at Home in Healthy Subjects and Fibromyalgia Patients. J Vis Exp. 2018 Jul 14;(137):57614. doi: 10.3791/57614. PMID: 30059026.

18. Barry RJ, Clarke AR, Johnstone SJ, Magee CA, Rushby JA. EEG differences between eyes-closed and eyes-open resting conditions. Clin Neurophysiol. 2007 Dec;118(12):2765–73. doi: 10.1016/j.clinph.2007.07.028. PMID: 17911042.

19. Barry RJ, De Blasio FM, Fogarty JS, Clarke AR. Natural alpha frequency components in resting EEG and their relation to arousal. Clin Neurophysiol. 2020 Jan;131(1):205–212. doi: 10.1016/j.clinph.2019.10.018. PMID: 31812081.

20. Delorme A, Makeig S. EEGLAB: an open source toolbox for analysis of single-trial EEG dynamics including independent component analysis. J Neurosci Methods. 2004 Mar 15;134(1):9–21. doi: 10.1016/j.jneumeth.2003.10.009. PMID: 15102499.

21. Ponomarev VA, Mueller A, Candrian G, Grin-Yatsenko VA, Kropotov JD. Group Independent Component Analysis (gICA) and Current Source Density (CSD) in the study of EEG in ADHD adults. Clin Neurophysiol. 2014 Jan;125(1):83–97. doi: 10.1016/j.clinph.2013.06.015. PMID: 23871197.

22. Jäncke L, Alahmadi N. Resting State EEG in Children With Learning Disabilities: An Independent Component Analysis Approach. Clin EEG Neurosci. 2016 Jan;47(1):24–36. doi: 10.1177/1550059415612622. PMID: 26545819.

23. Pascual-Marqui RD, Michel CM, Lehmann D. Low resolution electromagnetic tomography: a new method for localizing electrical activity in the brain. Int J Psychophysiol. 1994 Oct;18(1):49–65. doi: 10.1016/0167-8760(84)90014-x. PMID: 7876038.

24. Pascual-Marqui RD. Standardized low-resolution brain electromagnetic tomography (sLORETA): technical details. Methods Find Exp Clin Pharmacol. 2002;24 Suppl D:5-12. PMID: 12575463.

25. Pascual-Marqui, RD. Instantaneous and lagged measurements of linear and nonlinear dependence between groups of multivariate time series: frequency decomposition. arXiv:0711.1455 [Preprint]. 2007. Available from: https://arxiv.org/abs/0711.1455

26. Mazziotta J, Toga A, Evans A, Fox P, Lancaster J, Zilles K, et al. A probabilistic atlas and reference system for the human brain: International Consortium for Brain Mapping (ICBM). Philos Trans R Soc Lond B Biol Sci. 2001 Aug 29;356(1412):1293–322. doi: 10.1098/rstb.2001.0915. PMID: 11545704.

27. Fuchs M, Kastner J, Wagner M, Hawes S, Ebersole JS. A standardized boundary element method volume conductor model. Clin Neurophysiol. 2002 May;113(5):702–12. doi: 10.1016/s1388-2457(02)00030-5. PMID: 11976050.

28. Jurcak V, Tsuzuki D, Dan I. 10/20, 10/10, and 10/5 systems revisited: their validity as relative head-surface-based positioning systems. Neuroimage. 2007 Feb 15;34(4):1600–11. doi: 10.1016/j.neuroimage.2006.09.024. PMID: 17207640.

29. Pascual-Marqui, RD. Coherence and phase synchronization: generalization to pairs of multivariate time series, and removal of zero-lag contributions. arXiv:0706.1776v3 [Preprint]. 2007. Available from: http://arxiv.org/abs/0706.1776

30. Coben R, Mohammad-Rezazadeh I, Cannon RL. Using quantitative and analytic EEG methods in the understanding of connectivity in autism spectrum disorders: a theory of mixed over- and under-connectivity. Front Hum Neurosci. 2014 Feb 26;8:45. doi: 10.3389/fnhum.2014.00045. PMID: 24616679.

31. Bosch-Bayard J, Biscay RJ, Fernandez T, Otero GA, Ricardo-Garcell J, Aubert-Vazquez E, et al. EEG effective connectivity during the first year of life mirrors brain synaptogenesis, myelination, and early right hemisphere predominance. Neuroimage. 2022 May 15;252:119035. doi: 10.1016/j.neuroimage.2022.119035. PMID: 35218932.

32. Dai YJ, Zhang X, Yang Y, Nan HY, Yu Y, Sun Q, et al. Gender differences in functional connectivities between insular subdivisions and selective pain-related brain structures. J Headache Pain. 2018 Mar 14;19(1):24. doi: 10.1186/s10194-018-0849-z. PMID: 29541875.

33. Wang Z, Yuan M, Xiao J, Chen L, Guo X, Dou Y, et al. Gray Matter Abnormalities in Patients with Chronic Primary Pain: A Coordinate-Based Meta-Analysis. Pain Physician. 2022 Jan;25(1):1–13. PMID: 35051138.

34. Cifre I, Sitges C, Fraiman D, Muñoz MÁ, Balenzuela P, González-Roldán A, et al. Disrupted functional connectivity of the pain network in fibromyalgia. Psychosom Med. 2012 Jan;74(1):55–62. doi: 10.1097/PSY.0b013e3182408f04. PMID: 22210242.

35. Fauchon C, Meunier D, Faillenot I, Pomares FB, Bastuji H, Garcia-Larrea L, et al. The Modular Organization of Pain Brain Networks: An fMRI Graph Analysis Informed by Intracranial EEG. Cereb Cortex Commun. 2020 Nov 25;1(1):tgaa088. doi: 10.1093/texcom/tgaa088. PMID: 34296144.

36. Marques AP, Santos AMB, Assumpção A, Matsutani LA, Lage LV, Pereira CAB. Validação da versão brasileira do Fibromyalgia Impact Questionnaire (FIQ). Rev Bras Reumatol Engl Ed. 2006; 46(1), 24–31.

37. Gomes-Oliveira MH, Gorenstein C, Lotufo Neto F, Andrade LH, Wang YP. Validation of the Brazilian Portuguese version of the Beck Depression Inventory-II in a community sample. Braz J Psychiatry. 2012 Dec;34(4):389–94. doi: 10.1016/j.rbp.2012.03.005. PMID: 23429809.

38. Sehn F, Chachamovich E, Vidor LP, Dall-Agnol L, de Souza IC, Torres IL, et al. Cross-cultural adaptation and validation of the Brazilian Portuguese version of the pain catastrophizing scale. Pain Med. 2012 Nov;13(11):1425–35. doi: 10.1111/j.1526-4637.2012.01492.x. PMID: 23036076.

39. Bertolazi AN, Fagondes SC, Hoff LS, Dartora EG, Miozzo IC, de Barba ME, et al. Validation of the Brazilian Portuguese version of the Pittsburgh Sleep Quality Index. Sleep Med. 2011 Jan;12(1):70–5. doi: 10.1016/j.sleep.2010.04.020. PMID: 21145786.

40. Amorim P. Mini International Neuropsychiatric Interview (MINI): validação de entrevista breve para diagnóstico de transtornos mentais. Rev Bras Psiquiatr. 2000; 22 (3), 06–115.

41. Caumo W, Antunes LC, Elkfury JL, Herbstrith EG, Busanello Sipmann R, et al. The Central Sensitization Inventory validated and adapted for a Brazilian population: psychometric properties and its relationship with brain-derived neurotrophic factor. J Pain Res. 2017 Sep 1;10:2109–2122. doi: 10.2147/JPR.S131479. PMID: 28979158.

42. Galambos A, Szabó E, Nagy Z, Édes AE, Kocsel N, Juhász G, et al. A systematic review of structural and functional MRI studies on pain catastrophizing. J Pain Res. 2019 Apr 11;12:1155–1178. doi: 10.2147/JPR.S192246. PMID: 31114299.

43. Harris RE, Sundgren PC, Craig AD, Kirshenbaum E, Sen A, Napadow V, et al. Elevated insular glutamate in fibromyalgia is associated with experimental pain. Arthritis Rheum. 2009 Oct;60(10):3146–52. doi: 10.1002/art.24849. PMID: 19790053.

44. Čeko M, Shir Y, Ouellet JA, Ware MA, Stone LS, Seminowicz DA. Partial recovery of abnormal insula and dorsolateral prefrontal connectivity to cognitive networks in chronic low back pain after treatment. Hum Brain Mapp. 2015 Jun;36(6):2075–92. doi: 10.1002/hbm.22757. Epub 2015 Feb 3. PMID: 25648842.

45. Seminowicz DA, Moayedi M. The Dorsolateral Prefrontal Cortex in Acute and Chronic Pain. J Pain. 2017 Sep;18(9):1027–1035. doi: 10.1016/j.jpain.2017.03.008. Epub 2017 Apr 8. PMID: 28400293.

46. Pizzagalli DA. Frontocingulate dysfunction in depression: toward biomarkers of treatment response. Neuropsychopharmacology. 2011 Jan;36(1):183–206. doi: 10.1038/npp.2010.166. Epub 2010 Sep 22. PMID: 20861828.

47. Dennis TA, Solomon B. Frontal EEG and emotion regulation: electrocortical activity in response to emotional film clips is associated with reduced mood induction and attention interference effects. Biol Psychol. 2010 Dec;85(3):456–64. doi: 10.1016/j.biopsycho.2010.09.008. PMID: 20863872; PMCID: PMC2976487.

48. Baliki MN, Mansour AR, Baria AT, Apkarian AV. Functional reorganization of the default mode network across chronic pain conditions. PLoS One. 2014 Sep 2;9(9):e106133. doi: 10.1371/journal.pone.0106133. PMID: 25180885.

49. Pizzagalli DA, Roberts AC. Prefrontal cortex and depression. Neuropsychopharmacology. 2022 Jan;47(1):225–246. doi: 10.1038/s41386-021-01101-7. Epub 2021 Aug 2. Erratum in: Neuropsychopharmacology. 2021 Aug 19;: PMID: 34341498.

50. Cao H, Gao YJ, Ren WH, Li TT, Duan KZ, Cui YH, et al. Activation of extracellular signal-regulated kinase in the anterior cingulate cortex contributes to the induction and expression of affective pain. J Neurosci. 2009 Mar 11;29(10):3307–21. doi: 10.1523/JNEUROSCI.4300-08.2009. PMID: 19279268.

51. Bliss TV, Collingridge GL, Kaang BK, Zhuo M. Synaptic plasticity in the anterior cingulate cortex in acute and chronic pain. Nat Rev Neurosci. 2016 Aug;17(8):485–96. doi: 10.1038/nrn.2016.68. PMID: 27307118.

52. Fregni F, El-Hagrassy MM, Pacheco-Barrios K, Carvalho S, Leite J, Simis M, et al. Evidence-Based Guidelines and Secondary Meta-Analysis for the Use of Transcranial Direct Current Stimulation in Neurological and Psychiatric Disorders. Int J Neuropsychopharmacol. 2021 Apr 21;24(4):256–313. doi: 10.1093/ijnp/pyaa051. PMID: 32710772.

53. Harmony T. The functional significance of delta oscillations in cognitive processing. Front Integr Neurosci. 2013 Dec 5;7:83. doi: 10.3389/fnint.2013.00083. PMID: 24367301.

54. Xiao X, Ding M, Zhang YQ. Role of the Anterior Cingulate Cortex in Translational Pain Research. Neurosci Bull. 2021 Mar;37(3):405–422. doi: 10.1007/s12264-020-00615-2. Epub 2021 Feb 10. PMID: 33566301.

55. Mariano TY, Burgess FW, Bowker M, Kirschner J, Van’t Wout-Frank M, et al. Transcranial Direct Current Stimulation for Affective Symptoms and Functioning in Chronic Low Back Pain: A Pilot Double-Blinded, Randomized, Placebo-Controlled Trial. Pain Med. 2019 Jun 1;20(6):1166–1177. doi: 10.1093/pm/pny188. PMID: 30358864.

56. Kim J, Loggia ML, Cahalan CM, Harris RE, Beissner F Dr Phil Nat, Garcia RG, et al. The somatosensory link in fibromyalgia: functional connectivity of the primary somatosensory cortex is altered by sustained pain and is associated with clinical/autonomic dysfunction. Arthritis Rheumatol. 2015 May;67(5):1395–1405. doi: 10.1002/art.39043. PMID: 25622796.

57. Kim J, Mawla I, Kong J, Lee J, Gerber J, Ortiz A, et al. Somatotopically specific primary somatosensory connectivity to salience and default mode networks encodes clinical pain. Pain. 2019 Jul;160(7):1594–1605. doi: 10.1097/j.pain.0000000000001541. PMID: 30839429.

58. Zortea M, Beltran G, Alves RL, Vicuña P, Torres ILS, Fregni F, et al. Spectral Power Density analysis of the resting-state as a marker of the central effects of opioid use in fibromyalgia. Sci Rep. 2021 Nov 22;11(1):22716. doi: 10.1038/s41598-021-01982-0. PMID: 34811404.

59. Dumville JC, Hahn S, Miles JN, Torgerson DJ. The use of unequal randomisation ratios in clinical trials: a review. Contemp Clin Trials. 2006 Feb;27(1):1–12. doi: 10.1016/j.cct.2005.08.003. PMID: 16236557.

60. Hey SP, Kimmelman J. The questionable use of unequal allocation in confirmatory trials. Neurology. 2014 Jan 7;82(1):77–9. doi: 10.1212/01.wnl.0000438226.10353.1c. Epub 2013 Dec 4. PMID: 24306005.

61. Michel CM, Brunet D. EEG Source Imaging: A Practical Review of the Analysis Steps. Front Neurol. 2019 Apr 4;10:325. doi: 10.3389/fneur.2019.00325. PMID: 31019487.

62. Painold A, Faber PL, Reininghaus EZ, Mörkl S, Holl AK, Achermann P, et al. Reduced Brain Electric Activity and Functional Connectivity in Bipolar Euthymia: An sLORETA Source Localization Study. Clin EEG Neurosci. 2020 May;51(3):155–166. doi: 10.1177/1550059419893472. Epub 2019 Dec 17. PMID: 31845595.

63. Yoshimura M, Pascual-Marqui R, Nishida K, Kitaura Y, Mii H, Saito Y. P2-3-8 Change of cross frequency coupling by symptom provocation in Obsessive Compulsive Disorder (OCD) based on sLORETA. Clin Neurophysiol. 2018; 29(5): e39. doi: 10.1016/j.clinph.2018.02.103

64. Vanneste S, Heyning PV, Ridder DD. Contralateral parahippocampal gamma-band activity determines noise-like tinnitus laterality: a region of interest analysis. Neuroscience. 2011 Dec 29;199:481–90. doi: 10.1016/j.neuroscience.2011.07.067. PMID: 21920411.

65. Vanneste S, Ost J, Van Havenbergh T, De Ridder D. Resting state electrical brain activity and connectivity in fibromyalgia. PLoS One. 2017 Jun 26;12(6):e0178516. doi: 10.1371/journal.pone.0178516. PMID: 28650974.

66. Russell MJ, Goodman TA, Visse JM, Beckett L, Saito N, Lyeth BG, Recanzone GH. Sex and Electrode Configuration in Transcranial Electrical Stimulation. Front Psychiatry. 2017 Aug 14;8:147. doi: 10.3389/fpsyt.2017.00147. PMID: 28855877.

67. Kuo MF, Paulus W, Nitsche MA. Sex differences in cortical neuroplasticity in humans. Neuroreport. 2006 Nov 6;17(16):1703–7. doi: 10.1097/01.wnr.0000239955.68319.c2. PMID: 17047457.

